# Logistic mixed-effects model analysis with pseudo-observations for estimating risk ratios in clustered binary data analysis

**DOI:** 10.1101/2025.06.26.25330352

**Authors:** Hisashi Noma, Masahiko Gosho

## Abstract

Logistic mixed-effects model has been a standard multivariate analysis method for analyzing clustered binary outcome data, e.g., longitudinal studies, clustered randomized trials, and multi-center/regional studies. However, the resultant odds ratio estimator cannot be directly interpreted as an effect measure, and it is only interpreted as an approximation of the risk ratio estimator when the frequency of events is small. In this article, we propose a new statistical analysis method that enables to provide risk ratio estimator in the multilevel statistical model framework. The valid risk ratio estimation is realized via augmenting pseudo-observations to the original dataset and then analyzing the modified dataset by the logistic mixed-effects model. The resultant estimators of fixed effect coefficients are theoretically shown to be consistent estimators of the risk ratios. Also, the standard errors and confidence intervals of the risk ratios can be calculated by bootstrap method. All of the computations are simply implementable by using R package “glmmrr”. We illustrate the effectiveness of the proposed method via applications to a cluster-randomized trial of maternal and child health handbook and a longitudinal study of respiratory disease. Also, we provide simulation-based evidence for the accuracy and precision of estimation of risk ratios by the proposed method.

## 1. Introduction

Logistic mixed-effects model has been a standard multivariate analysis method for analyzing clustered binary outcome data in clinical and epidemiological studies, e.g., longitudinal studies, clustered randomized trials, and multi-center/regional studies ^1-3^. However, the resultant odds ratio estimator cannot be directly interpreted as an effect measure, and it is only interpreted as an approximation of the risk ratio estimator when the frequency of events is small ^4,5^. Similar problems have been widely discussed for unclustered data analyses, e.g., the modified Poisson regression ^6^ is one of the representative methods that enables unbiased estimation of risk ratios in the multivariate analyses. The Poisson regression approach can be extended to the generalized estimating equation (GEE) framework for analyzing clustered binary data ^7^, but the estimating equation-based semiparametric inference method cannot be straightforwardly extended to the fully parametric multilevel modelling framework. A remarkable advantage of the multilevel modelling is that can be implemented by the direct likelihood principle for analysis of incomplete data ^1,8^; the influence of missing data is ignorable if the missing data mechanism is missing-at-random (MAR) ^9^. For this reason, the mixed-effects models are also widely used for primary analyses of longitudinal clinical trials ^10^. The ignorability is not fulfilled for the GEE approach, and currently, no effective statistical methods are available for estimating risk ratios directly on the multilevel modelling framework.

In this article, we propose a new effective method in estimating risk ratios for clustered binary outcome data in the multilevel modelling framework. Especially, we theoretically show the regression coefficient estimates obtained from a logistic mixed-effects model fitting to a modified dataset, that some pseudo-observations are augmented, can be interpreted as unbiased estimates of risk ratios. This method can be simply implementable using standard statistical packages of the ordinary logistic mixed-effects model, and the confidence intervals of risk ratios can be calculated by bootstrap method. In addition, the Little-Rubin’s ignorability for missing data ^9^ is fulfilled for the proposed method. We illustrate the effectiveness of the proposed method via applications to a cluster-randomized trial of maternal and child health handbook ^11^ and a longitudinal clinical study of respiratory disease ^12^. We also provide simulation-based evidence for the accuracy and precision of estimation of risk ratios by the proposed method.

## 2. The pseudo-observations approach

### 2.1 Target statistical model and estimands

We consider *Y*_*ij*_ as the jth binary outcome (= 1: event occurred, = 0: event did not occur) on cluster *i*, where *i* = 1, …, N, and *j* = 1, …, *n*_*i*_. For instance, *Y*_*ij*_ is a repeated binary outcome on *j* th visit of *i* th individual for a longitudinal study, or outcome variable of jth participant of *i*th cluster for a cluster-randomized trial. Without loss of generality, we denote the number of events in cluster *i* as m_*i*_ and the total number of events throughout the *N* clusters as *M*. For analyzing the clustered binary data, the logistic mixed-effects model is one of the most widely used model in the generalized linear mixed-effects model (GLMM) that uses the canonical link of binomial outcome data ^8,13^,

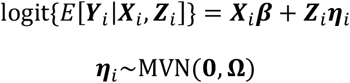

where ***X***_*i*_ and ***Z***_*i*_ are *n*_*i*_ × *p* and *n*_*i*_ × *q* design matrices, ***β*** is the *p* × 1 vector containing the fixed effects, and ***η***_*i*_ is the *q* × 1 vector containing the random effects; ***η***_*i*_ is assumed to be normally distributed with mean zero and covariance matrix *Ω* (*q* × *q* symmetric matrix). Conditional on the random effects ***η***_*i*_, the responses 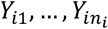 are independent and follow a multivariate Bernoulli distribution with a mean vector *E*[***Y***_*i*_|***X***_*i*_, ***Z***_*i*_]. The regression coefficients of *β* and the covariance matrix Ω are estimated by the (restricted) maximum likelihood (ML) method, and the resultant estimators are consistent and the most precise ones under large sample settings ^8,13^. The computations are implementable using standard statistical packages of GLMM, e.g., lme4 package ^14^ of R (R Foundation for Statistical Computing, Vienna, Austria), PLOC GLIMMIX of SAS (SAS Institute, Inc, Cary, NC). Also, potential influences from missing data are ignorable owing to the direct likelihood principle ^8,9^. However, the regression coefficients of ***β*** are interpreted as log-transformed odds-ratios as like the ordinary logistic regression, and only be interpreted as approximations of log risk ratios when the event frequency is small ^4,5^.

To overcome the interpretability issue, an alternative mixed-effects model that is based on the log-linear-type binomial mixed-effects model can be considered,

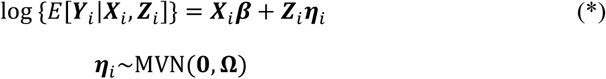

that is an extension of the log-linear binomial regression model ^13^ to the GLMM framework. The marginal regression model of jth component of ***Y***_*i*_ is expressed as

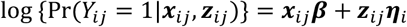

where ***x***_*ij*_ and ***z***_*ij*_ are the jth row vectors of ***X***_*i*_ and ***Z***_*i*_. The log-linear binomial regression model has a substantial computational problem because the values of the regression function can become > 1, and the ML estimates of regression coefficients cannot be obtained in many cases ^15,16^. The GLMM model also has the same computational problem, and the ML estimates cannot be often calculated. To resolve the computational issue of the log-linear binomial regression model, the modified Poisson regression ^6^ is developed based on the semiparametric estimating equation theory ^17,18^. However, the Poisson regression approach cannot be straightforwardly generalized to the fully parametric mixed-effects model, because the variance function is misspecified. In the following sections, we will show an alternative pseudo-likelihood method can resolve this issue, and we can provide a consistent estimator of ***β*** in the log-linear mixed-effects model (*) via simple computations by the ordinary GLMM software packages. Note that the following discussions assume the random-effects distribution is the multivariate normal distribution, it can be generalized to other flexible distributions ^19^. We formally adopt the standard assumption here that is used in most of current standard software packages, the following discussions can be extended to the other non-normal distribution models straightforwardly.

### 2.2. A pseudo-observations approach to provide risk-ratio estimator

The principle of the new method is adjusting the estimating function of the logistic mixed-effects model via adding pseudo-observations. The estimating algorithm for providing consistent risk ratio estimators are given as follows:

#### Algorithm 1 (Logistic mixed-effects model with pseudo-observations)

1. For the clustered binary outcome dataset {*Y*_*ij*_, ***x***_*ij*_***z***_*ij*_} (i = 1, … *N*; *j* = 1, …, *n*_*i*_),, create pseudo-observations for all of the individual units with *Y*_*ij*_ = 1 that have the same cluster indicators, ***x***_*ij*_, ***z***_*ij*_, and the outcomes are 0. Then, add the total *M* pseudo-observations to the original dataset; we denote the pseudo-observations as {*Y*_*ij*_, ***x***_*ij*_***z***_*ij*_} (*i* = 1, …, *N*; *j*= *n*_*i*_ + 1, …, *n_i_* + *m*_*i*_), which the order of running index j is anonymous.
2. Fit the logistic mixed-effects model to the augmented dataset that the pseudo-observations are added. The resultant estimators of regression coefficients and covariance parameters in {***β***,***Ω***} of the logistic mixed-effects model are consistent estimators of those of the log-linear mixed effects models (*), and the exponential-transformed regression coefficients estimators exp 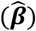 can be interpreted as consistent risk ratio estimators.

A remarkable advantage of this estimating method is the simplicity of the computation. It can be realized by simply fitting computational functions of the logistic mixed-effects model to the augmented dataset. The unbiasedness under practical situations are assessed in simulation studies in Section 4. Also, details of the mathematical proof of the consistency of the resultant estimators are presented in Appendix. Note the copy data with event indicators 0 are formally added, and we should regard them as pseudo-observations (not actually exist) that have statistical information corresponding to augmented likelihoods. The basic principle of the validity of the risk ratio estimation is founded on the estimating equation theory of the conventional case-control and case-cohort designs ^20-23^.

To estimate the standard errors of the resultant pseudo-likelihood estimators, the sandwich variance estimator would be a straightforward choice because of the estimating equation theory ^24^. However, the augmented data involves artificial duplicated data, that do not actually exist, thus the ordinary estimating equation theory does not work for this case ^23^. As shown in the simulation studies in Section 4, the sandwich variance estimator has serious bias even under large sample settings. However, we can quantify the statistical errors simply adapting the bootstrap method to the clustered data. Various parametric and nonparametric bootstrap approaches have been proposed to the GLMM framework ^25^, and as shown in the simulation studies in Section 4, the most common cluster-based nonparametric bootstrap could provide stable and valid inference results generally.

#### Algorithm 2 (Nonparametric bootstrap for the pseudo-observations method)

1. Resample N clusters from the original dataset with replacement, and re-label the cluster indicators in the resampled dataset as 1’(*b*), 2’(*b*), …, N’(*b*) (b = 1, …, B;).
2. For the resampled N cluster data, create the pseudo-observations as described in Algorithm 1. Then, fit the logistic mixed-effects model to the augmented dataset. The resultant estimates 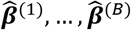 and 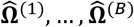 are the bootstrap samples of the pseudo-likelihood estimators of 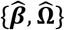

Using the bootstrap samples, confidence intervals of the elements of {***β, Ω***} can also be calculated, e.g., the 2.5th and 97.5th percentiles in the bootstrap samples can be used for calculating 95% confidence limits. The bootstrap P-values for statistical testing can also be calculable from the resampled data. The nonparametric bootstrap can effectively circumvent the parametric distribution assumptions and can provide stable inference results generally ^26^. For more complicated multilevel models involving non-nested hierarchical sampling models, parametric bootstrap techniques are also applicable ^25^. Note the number of resampling *B* should be ≥ 1,000 for providing stable confidence limits and P-values, generally. The logistic mixed-effects model requires numerical integrations for calculating ML estimates, and the bootstrap process might require large computational efforts, but under the recent computational environment, it is not realistically problematic; the parallel computation techniques can be effectively applied (as provided in the example R code in the Supporting Information materials).

### 2.3 Ignorability on missing data analysis

For the pseudo-observations approach, the Little-Rubin’s ignorability on missing data analysis ^9^ is straightforwardly fulfilled. We use a different notation in this section for convenience. Let *D* denote the set of complete data in the modified dataset that the pseudo-observations are added, and *R* a set of indicator variables indicating whether individual components of *D* are observed (=1) or missing (=0). Also, let *D* = (*D*_obs_, *D*_mis_), where *D*_obs_ is the observed part of *D* and *D*_mis_ is the missing part of *D*. Then, the joint probability functions of *D* and *R* for the logistic mixed-effects model is expressed as

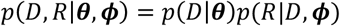

where ***θ*** is the parameter set of the data model and ***𝝓*** is that of the missing data mechanism model. Then the fully pseudo-likelihood function based on the observed data is

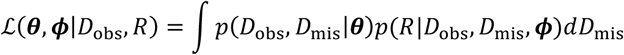

The MAR mechanism assumes the independency between *D*_mis_ and the missing data mechanism, i.e., *p*(*R*|*D*_obs_, *D*_mis_, ***𝝓***) = *p*(*R*|*D*_obs_, 𝝓), and the observed data pseudo-likelihood is expressed as

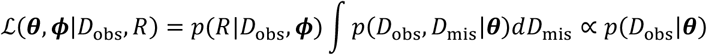

Thus, the missing data mechanism *p*(*R*|*D*_obs_, *𝝓*) is ignorable and we can conduct valid statistical inference for θ using the observed data pseudo-likelihood function under the MAR assumption ^9,27,28^. This theoretical property indicates that the proposed modified logistic mixed-effects model analysis can be used for incomplete data analysis as like the ordinary likelihood-based analyses using the mixed-effects models ^8,10^.

### 2.4 Software

We developed an R package, `glmmrr` (https://github.com/nomahi/glmmrr/), to perform the proposed methods via simple commands. Example R code for implementing these methods is provided in the Supporting Information materials.

## 3. Applications

### 3.1 Cluster-randomized trial of maternal and child health handbook

Mori et al. ^11^ conducted a cluster-randomized trial to assess the effectiveness of maternal and child health handbook in Mongolia. They randomly allocated the uses of the maternal and child health handbook to pregnant women in 18 districts, and assessed the effectiveness in promoting antenatal care attendance, especially whether the number of antenatal care visits were ≥ 6 times or not. 252 women in 9 districts were assigned to the intervention group, and 248 women in 9 districts were assigned to the control group. To assess the intervention effect, the intra-cluster correlations should be adequately adjusted, and the multilevel models have been one of the primary approaches ^2^. In assessing the intervention effect, the logistic mixed-effects models are the standard approach for analyzing the clustered binary outcome data, but the resultant odds ratio estimates are not interpretable effect measures if the frequency of events were not small ^4,5^. The crude event frequencies were 81.7% for the intervention group and 70.6% for the control group, and the logistic mixed-effects models would not be an adequate method for this case. In Table 1, we provide the odds ratio estimate by the logistic mixed-effects model involving random intercepts corresponding to 18 districts (the heterogeneity variance was denoted as τ^2^); the odds ratio estimate was 2.42 (95% confidence interval [CI]: 0.45, 13.10). The point estimate of the odds ratio indicated a large intervention effect, but due to the large event frequency, it could overestimate the actual relative risk, at least it was markedly large compared with the ratio of crude event frequencies. Mori et al. ^11^ also performed subgroup analyses by a summary index that quantified social-economic status of individual participants by the Demographic and Health Survey guidelines ^29^. They divided the population into 5 subgroups by quartiles of the social-economic index. We also provide the subgroup analysis results in Table 1. The effect sizes were especially large for the 1st and 2nd quartile subgroups (the richest 2 subgroups), and the odds ratio estimates were 7.48 (95%CI: 1.30, 42.97) and 35.25 (95%CI: 1.18, 1048.65), respectively. Besides, for the remaining subgroups, the intervention effects were not clearly shown, but the odds-ratio estimates themselves were certainly large (1.76, 1.55, and 0.77). These subgroup analyses indicated the effect modifications by the social-economic status; the impact of the maternal and child health handbook was larger for relatively wealthy and well-educated participants. However, the odds-ratio estimates could not quantify the intervention effects adequately.

**Table 1.** Results of the ordinary logistic mixed-effects model analysis and the pseudo-observations approach for the cluster randomized trial of maternal and child health handbook (*N* = 500).

|  | Logistic mixed-effects model |  | Pseudo-observations approach |  |
| --- | --- | --- | --- | --- |
| | OR estimates (95%CI) | $\tau$ estimate | RR estimates (95%CI) | $\tau$ estimate |
| All population | 2.42 (0.45, 13.10) | 1.66 | 1.16 (0.85, 1.56) | 0.08 |
| Subgroups by the wealth index |  |  |  |  |
| 1st quartile | 7.48 (1.30, 42.97) | 0.55 | 1.52 (0.99, 1.79) | 0.00 |
| 2nd quartile | 35.25 (1.18, 1048.65) | 2.06 | 1.52 (1.09, 4.71) | 0.00 |
| 3rd quartile | 1.76 (0.04, 71.62) | 3.26 | 1.05 (0.63, 1.63) | 0.00 |
| 4th quartile | 1.55 (0.22, 11.04) | 1.38 | 1.02 (0.73, 1.53) | 0.00 |
| 5th quartile | 0.77 (0.06, 9.83) | 1.85 | 0.90 (0.53, 1.23) | 0.00 |

As an alternative strategy, we performed the same analyses using the modified logistic mixed-effects model analyses with pseudo-observations. We augmented the original datasets with the pseudo-observations and simply fitted the logistic mixed-effects models with random intercepts corresponding to 18 districts to the modified datasets. The confidence intervals were calculated by the bootstrap method with 1,000 resamplings. The resultant risk ratio estimates and 95%CIs are presented in Table 1. At first, the risk ratio estimate for the overall population was 1.16 (95%CI: 0.85, 1.56). The magnitude of relative risk estimate was quite small compared with the odds ratio estimate, but would be considered as plausible comparing the crude event frequencies of the two groups. For the subgroup analyses by the social-economic index, the risk ratio estimates were also moderate compared to the odds ratio estimates. For the 1st and 2nd quartile subgroups, the risk ratio estimates were 1.52 (95%CI: 0.99, 1.79) and 1.52 (95%CI: 1.09, 4.71) respectively, and there also were great discrepancies with the odds ratio estimates. Also, for the 3rd, 4th and 5th quartile subgroups, the risk ratio estimates were 1.05 (95%CI: 0.63, 1.63), 1.02 (95%CI: 0.73, 1.53) and 0.90 (95%CI: 0.53, 1.23); these estimates indicated quite small between-groups differences. Note the τ estimate of the pseudo-observations approach was much smaller than that of the ordinary logistic mixed-effects model, because the sizes of odds ratios are usually estimated much larger than those of risk ratios when the event rate is large ^3^; variability of the odds ratios was also largely estimated. All of these results indicated the odds-ratio estimates could provide misleading evidence that overestimated the actual relative risks for the high frequency events. The pseudo-observations approach would provide more precise information for the intervention effects.

### 3.2 Longitudinal clinical study for respiratory disease

Statistical analyses of longitudinal studies also require appropriate adjustments of correlations for repeated measurements, and the multilevel models are one of the primary statistical tools for them. Also, the direct likelihood or Bayesian approaches can address the influences of missing data by the MAR mechanism owing to the Little-Rubin’s ignorablity principle ^9^. Koch et al. ^12^ provided a longitudinal clinical trial data for respiratory disease, where 111 participants from two centers were randomly assigned to active treatment or placebo. The primary outcome was dichotomized respiratory status (good or poor), and the respiratory statuses were planned to be measured at baseline and four visits after randomization. The crude event frequencies were 68.1% for the active treatment group and 44.3% for the placebo group. We firstly fitted the logistic mixed-effects model with random intercepts corresponding to the individual participants (the heterogeneity variance was denoted as τ^2^) and involving treatment (active vs. placebo), baseline respiratory status, center, gender (male vs. female), and age as the fixed effects to the clinical study data. The results are presented in Table 2. The odds ratio estimate for the treatment was 7.55 (95%CI: 2.61, 21.84) and indicated a remarkable treatment effect. Also, that for the baseline symptom was 18.36 (95%CI: 5.92, 56.96) and indicated a strong prognostic effect. For the remaining variables, the 95%CIs of odds ratios covered the null value (=1), but the point estimates indicated certainly large prognostic effects.

**Table 2.** Results of the ordinary logistic mixed-effects model analysis and the pseudo-observations approach for the longitudinal clinical study of respiratory disease (*N* = 111).

|  | Logistic mixed-effects model | Pseudo-observations approach |
| --- | --- | --- |
|  | OR estimates (95%CI) | RR estimates (95%CI) |
| Treatment (ref: placebo) | 7.55 (2.61, 21.84) | 1.61 (1.22, 2.17) |
| Baseline symptom | 18.36 (5.92, 56.96) | 2.04 (1.56, 2.85) |
| Center | 2.66 (0.92, 7.71) | 1.32 (1.01, 1.75) |
| Gender (ref: female) | 0.78 (0.21, 2.92) | 0.95 (0.65, 1.44) |
| Age | 0.97 (0.94, 1.01) | 0.99 (0.98, 1.00) |
| $\tau$ estimate | 1.98 | 0.00 |

To assess the risk ratios directly, we fitted the logistic mixed-effects model augmented the pseudo-observations to the large event frequency data. The resultant risk ratio estimates and 95%CIs are presented in Table 2. The risk ratio estimate of the treatment was 1.61 (95%CI: 1.22, 2.17); significant treatment effect was indicated, but the magnitude of the effect measure estimate was much smaller than that of the odds ratio. Also, the risk ratio estimate of the baseline symptom was 2.04 (95%CI: 1.56, 2.85), and the magnitudes of size of the estimate were quite different. For the remaining variables, all of the risk ratio estimates became moderate. For the longitudinal study with high frequency events, the overall results and interpretations would be entirely changed through using the risk ratio estimation method.

## 4. Simulation studies

To illustrate the operating characteristics of the proposed method, we performed simulation studies. For data generation, to replicate the correlation structures of real-world data, we resampled the participant level data from the longitudinal clinical study of respiratory disease in the previous section. Then, we supposed the binomial log-linear mixed effects model (*) for the outcome data generation,

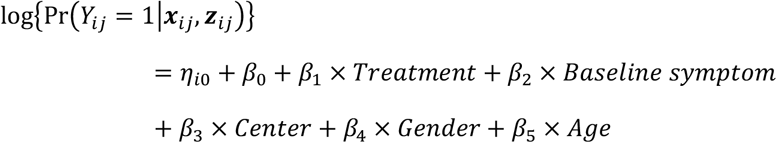

where *η*_i0_~*N*(0, *τ*^2^) was the random intercept and ***β*** = (*β*_1_, …, *β*_5_)*T* was the fixed effect regression coefficients (*i* = 1, …, *N*; *j* = 1, …,4). We considered several scenarios that chanted *N* = 50, 100, 200 and 500, and τ = 0.02, 0.05, and 0.10. Also, the log risk ratio *β*_1_ that expressed the treatment effect was set to 0.47 (= log(1.60)) and 0.69 (= log(2.00)); the other coefficients were set to the estimates obtained from the analysis of the original data, *β*_2_ = 0.71, *β*_3_ = 0.28, *β*_4_ = −0.05, and *β*_5_ = −0.01. *β*_0_ was set to an appropriate value so that the overall event frequency (P_event_) became 0.20 and 0.40. In addition, we considered a MAR dropout mechanism that follows logit{Pr(*R*_*ij*_ = 1|*Y*_*i*,*j*−1_)} = *𝜉*_0_ + log (1.50) × *Y*_*i*,*j*−1_, where *R*_*ij*_ is the indicator variable for the dropout at *j*th visit of the *i*th participant (*i* = 1, …, *N*; *j* = 2, …,4); *𝜉*_0_ is set such that the total dropout rate is 20%. We performed 1,000 simulations for each scenario, and the simulated datasets were analyzed by the ordinary logistic mixed-effects model and the proposed modified logistic mixed-effects model. For calculating bootstrap CIs, we performed 1,000 resampling consistently. For the evaluation measures, we calculated the means and standard deviations of the odds ratio and risk ratio estimates corresponding to *β*_1_ obtained from 1,000 simulations. Also, for the proposed method, we calculated mean of the standard error estimates by bootstrap and root mean squared error (RMSE) of the estimates of *β*_1_, and the coverage rate and expected width of 95% CIs of *β*_1_.

The simulation results are presented in Tables 3 and 4. For the ordinary logistic mixed-effects model analyses, the mean of the estimates of odds ratio for the treatment effect was seriously overestimated and biased from the true risk ratio, because of the large event frequencies. Besides, using the pseudo-observations approach, the risk ratios were accurately estimated under all scenarios. These results indicate that the ordinary logistic mixed-effects model could not provide interpretable effect measure estimates, and the pseudo-observations approach clearly improved this relevant issue. Also, the bootstrap standard error estimates for the proposed method accurately estimated the actual standard errors; the accuracy depended on the sample size, and it was relatively improved if the sample size became larger. The RMSEs were nearly equivalent to the standard errors of the log risk ratio estimates, because the biases were generally to be 0. For the bootstrap confidence intervals, the coverage rates were consistently around the nominal level (=0.95) and could provide valid interval estimates generally. The expected widths depended on the parameter settings, especially *N* and P_event_. Note that the standard error estimates could be biased under moderate *N* setting (i.e., *N* = 50), because the validity is founded on the large sample theory; but even under these settings, coverage rates of the confidence interval were favorable.

**Table 3.** Results of the simulation studies (I): Comparison of the ordinary logistic mixed-effects model and the pseudo-observations approach (N = 50, 100) ^†^.

| $N$ | $P_{\text{event}}$ | $\tau$ | $\beta_1$ | Linear mixed-effects model<br>(log odds ratio) | | Pseudo-observations approach (log risk ratio) | | | | | |
| --- | --- | --- | --- | --- | --- | --- | --- | --- | --- | --- | --- |
| | | | | Mean | SE | Mean | SE | $\widehat{\text{SE}}$ | RMSE | CR | EW |
| 50 | 0.20 | 0.02 | 0.47 | 0.664 | 0.467 | 0.483 | 0.351 | 0.450 | 0.351 | 0.943 | 1.531 |
| 50 | 0.20 | 0.02 | 0.69 | 1.003 | 0.444 | 0.731 | 0.335 | 0.484 | 0.338 | 0.957 | 1.622 |
| 50 | 0.20 | 0.05 | 0.47 | 0.667 | 0.460 | 0.484 | 0.346 | 0.446 | 0.347 | 0.937 | 1.527 |
| 50 | 0.20 | 0.05 | 0.69 | 0.997 | 0.481 | 0.721 | 0.360 | 0.477 | 0.361 | 0.937 | 1.589 |
| 50 | 0.20 | 0.10 | 0.47 | 0.680 | 0.464 | 0.492 | 0.348 | 0.444 | 0.349 | 0.937 | 1.509 |
| 50 | 0.20 | 0.10 | 0.69 | 0.981 | 0.462 | 0.709 | 0.349 | 0.468 | 0.349 | 0.948 | 1.577 |
| 50 | 0.40 | 0.02 | 0.47 | 1.061 | 0.411 | 0.493 | 0.202 | 0.217 | 0.203 | 0.944 | 0.851 |
| 50 | 0.40 | 0.02 | 0.69 | 1.593 | 0.441 | 0.700 | 0.208 | 0.220 | 0.208 | 0.938 | 0.860 |
| 50 | 0.40 | 0.05 | 0.47 | 1.025 | 0.404 | 0.476 | 0.203 | 0.217 | 0.203 | 0.949 | 0.850 |
| 50 | 0.40 | 0.05 | 0.69 | 1.606 | 0.447 | 0.703 | 0.201 | 0.218 | 0.201 | 0.950 | 0.853 |
| 50 | 0.40 | 0.10 | 0.47 | 1.044 | 0.422 | 0.480 | 0.207 | 0.218 | 0.208 | 0.938 | 0.853 |
| 50 | 0.40 | 0.10 | 0.69 | 1.570 | 0.438 | 0.695 | 0.203 | 0.223 | 0.203 | 0.943 | 0.874 |
| 100 | 0.20 | 0.02 | 0.47 | 0.644 | 0.311 | 0.472 | 0.237 | 0.233 | 0.236 | 0.938 | 0.915 |
| 100 | 0.20 | 0.02 | 0.69 | 0.956 | 0.313 | 0.696 | 0.232 | 0.237 | 0.232 | 0.939 | 0.927 |
| 100 | 0.20 | 0.05 | 0.47 | 0.657 | 0.301 | 0.482 | 0.227 | 0.234 | 0.227 | 0.948 | 0.919 |
| 100 | 0.20 | 0.05 | 0.69 | 0.966 | 0.306 | 0.703 | 0.229 | 0.238 | 0.229 | 0.944 | 0.934 |
| 100 | 0.20 | 0.10 | 0.47 | 0.662 | 0.321 | 0.484 | 0.243 | 0.235 | 0.243 | 0.934 | 0.922 |
| 100 | 0.20 | 0.10 | 0.69 | 0.970 | 0.309 | 0.705 | 0.231 | 0.238 | 0.231 | 0.946 | 0.933 |
| 100 | 0.40 | 0.02 | 0.47 | 1.013 | 0.288 | 0.474 | 0.143 | 0.139 | 0.143 | 0.935 | 0.543 |
| 100 | 0.40 | 0.02 | 0.69 | 1.545 | 0.292 | 0.691 | 0.138 | 0.141 | 0.138 | 0.944 | 0.550 |
| 100 | 0.40 | 0.05 | 0.47 | 1.010 | 0.270 | 0.475 | 0.136 | 0.139 | 0.136 | 0.953 | 0.545 |
| 100 | 0.40 | 0.05 | 0.69 | 1.534 | 0.304 | 0.689 | 0.142 | 0.141 | 0.142 | 0.942 | 0.553 |
| 100 | 0.40 | 0.10 | 0.47 | 0.996 | 0.281 | 0.465 | 0.140 | 0.140 | 0.140 | 0.936 | 0.546 |
| 100 | 0.40 | 0.10 | 0.69 | 1.527 | 0.300 | 0.682 | 0.140 | 0.142 | 0.140 | 0.943 | 0.556 |

<sup>†</sup> Mean, SE: Mean and standard deviation of the estimates of $\beta_1$ , $\widehat{\text{SE}}$ : Mean of the standard error estimates of $\beta_1$ , RMSE: Root mean squared error of the estimates of $\beta_1$ , CR, EW: Coverage rate and expected width of the 95% confidence intervals of $\beta_1$ .

**Table 4.**
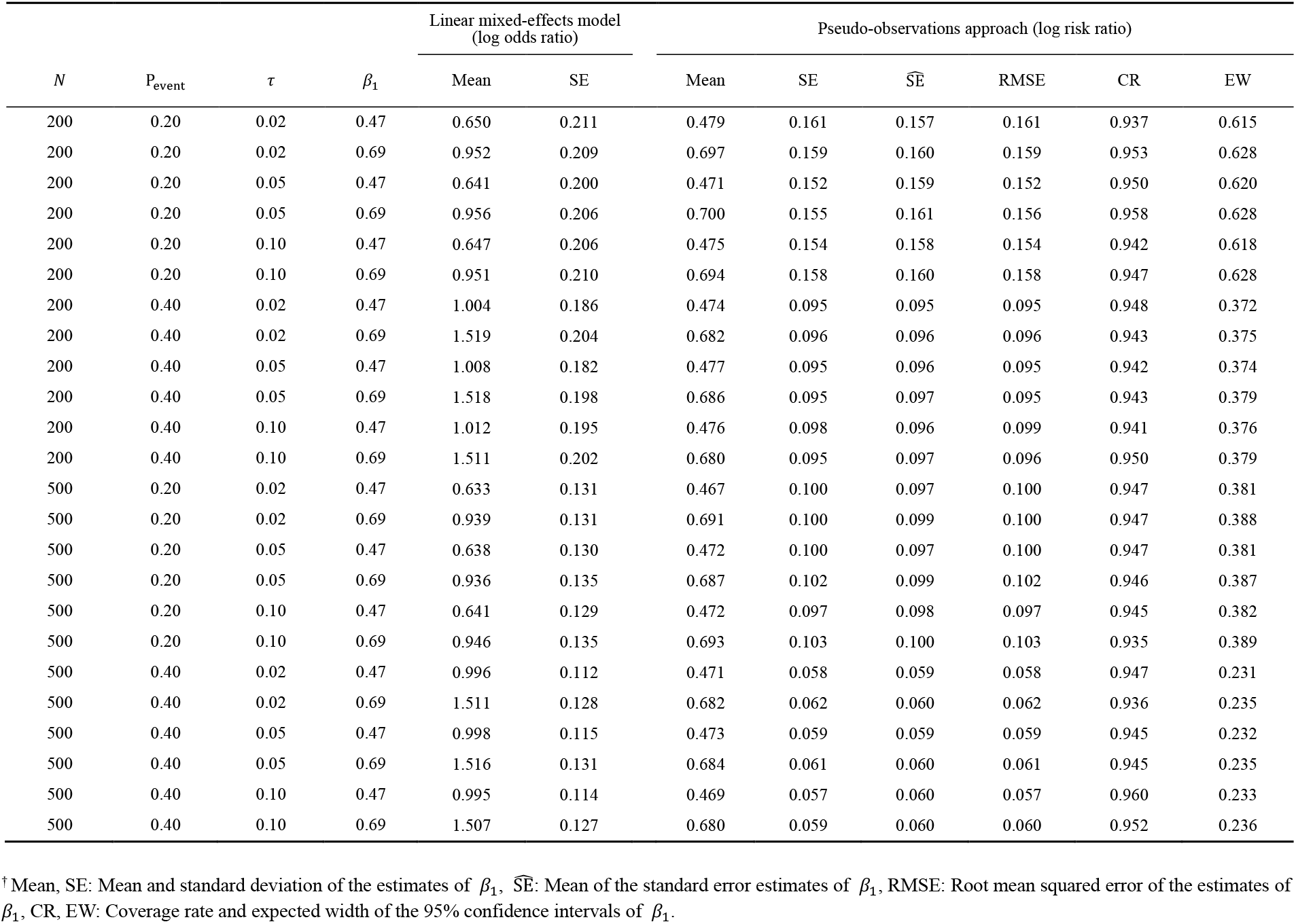
Results of the simulation studies (II): Comparison of the ordinary logistic mixed-effects model and the pseudo-observations approach (N = 200, 500) ^†^.

## 5. Discussion

Multilevel statistical modelling has been one of the standard statistical tools in clinical and epidemiological studies ^1-3^. However, the resultant effect measure estimates directly influence to the interpretation of the evidence and subsequent decision making for public health. The logistic mixed-effects model is mathematically convenient model that uses the canonical link function of binomial distribution ^13^, but the resultant odds ratio estimates have difficulties in the interpretations as shown in the numerical studies. Recently, for non-clustered data analyses, to overcome the practical issue of odds ratio, the modified Poisson regression ^6^ have been widely adopted in practice that can provide risk ratio estimates directly. However, similar effective solutions have not been proposed in the statistical analyses using multilevel statistical models. The pseudo-observations approach would provide effective and realistic solutions for this issue preserving the nice property of the Little-Rubin’s ignorability for missing data ^9^.

The practical advantage of the proposed methods is the computational conveniences. The pseudo-observation method can be implemented only adding pseudo-observations to the original dataset and then fitting the ordinary logistic mixed-effects model to the modified dataset. This process does not require special software packages and can be implemented using the standard GLMM packages. For computing confidence intervals, the bootstrap calculations are needed, but they can be performed by simple commands. The computational time is not generally so problematic, because parallel computation tools are available under current computational environments.

In addition, the risk ratio estimation methods are useful for more broad settings. For example, clinical trials for drug development often require correlated data analyses and the logistic mixed-effects model has been widely adopted, e.g., longitudinal clinical trials involving repeated outcome measurements ^10^, multicenter/regional clinical trials ^30^, and crossover trials ^31^. However, for these cases, the event frequencies are often not small, and the resultant odds ratio estimates have difficulty in measuring the magnitude of the treatment effects. The proposed risk ratio estimation method would also be effectively applied to these practical situations.

Similar ideas that modify the logistic regression analysis in ordinary binary outcome data analyses have been discussed in Dias-Quijano ^32^ and Dwivedi et al. ^33^. Our proposed method can be seen as the generalization of their approaches, but their confidence intervals depend on modified model variance estimates or sandwich variance estimates. The variance estimators can have serious biases ^23^ and bootstrapping approaches would be needed. Also, the generalization to the GEE framework of their methods would have difficulties, because the working correlation structures among the pseudo-observations and the other units in the same clusters are difficult to consider; additional theoretical assumptions are needed due to the artificial augmented data. In the GEE analysis setting, we consider the modified Poisson regression approach ^7^ would be more effective.

However, the Little-Rubin’s ignorablity for missing data ^9^ does not fulfill for the semiparametric method, and the pseudo-observations approach would have practical advantages in some aspects. Also, the GEE approach is used to estimate a population-averaged effect, while the multilevel modelling approach provides estimates of subject-specific effects ^34^; the estimands are essentially different between the two approaches and both options are relevant for practical applications. In addition, various advanced methods are developed for longitudinal data analyses, e.g., Bayesian modelling approaches ^35,36^, machine learning methods ^37^ and various sensitivity analysis methods for not MAR situations ^9,10,27^, and extensions of the proposed method to these frameworks would be important future issues.

In conclusion, the pseudo-observations approach using logistic mixed-effects model would provide new insights in clinical and epidemiological studies that require multilevel statistical models and would be an effective statistical tool for presenting more precise evidence for public health research and clinical practice.

## Data Availability

The original data used in this study is available in Mori et al. [11] and Kock et al. [12].

## Acknowledgements

This study was supported by Grants-in-Aid for Scientific Research from the Japan Society for the Promotion of Science (grant numbers: JP23K11931, JP22H03554, JP24K21306, and JP23H03063).

## Data availability statements

R package `glmmrr` for implementing the proposed methods is available at GitHub (https://github.com/nomahi/glmmrr/). Also, the original datasets are involved in the R package. Example R codes for replicating the results of the analyses are provided in the Supporting Information materials.

## Appendix Proof of the consistency of the pseudo-likelihood estimator

For the original dataset, we hypothetically conduct resampling without replacements: (a) from individual units (*i, j*) whose outcome is *Y*_*ij*_ = 1 with probability 1, and (b) from all individual units (*i, j*) with probability *λ*_0_. Then, for the resultant dataset composed of the selected units, we formally introduce indicator variables *D*_*ij*_ that value 1 for the former units and value 0 for the latter units. Note some units can be selected for both of the two resampling schemes duplicatedly, we formally regard them as different units in the selected units. Then, the conditional probability of *D*_*ij*_ = 1 given ***x***_*ij*_, ***z***_*ij*_ is expressed as

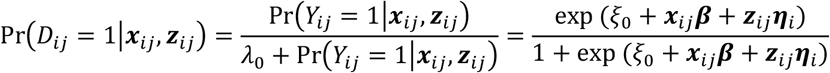

where *𝜉*_0_ = − log(*λ*_0_). This relationship shows the conditional probability of *D*_*ij*_ = 1 is expressed as the logistic transformation of *𝜉*_0_ + ***x***_*ij*_*β* + ***z***_*ij*_***η****_i_*. Besides, the conditional probability of *D*_*ij*_ = 0 is also expressed as

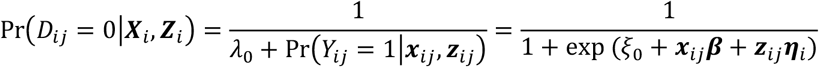

Then, through fitting the logistic mixed-effects model to the resampled dataset whose outcome variable is D_ij_ and setting the offset to *𝜉*_0_, we can obtain a consistent estimator of {***β, Ω***} of the log-linear mixed-effects model (*). As a special case of *λ*_0_ = 1, the resampling scheme exactly corresponds to the pseudo-observations augmentation in Algorithm 1, and then, *𝜉*_0_ = 0. Thus, the pseudo-likelihood estimator of **{*β, Ω***} obtained by Algorithm 1 is consistent estimators for the parameters in the log-linear mixed-effects model (*).

---

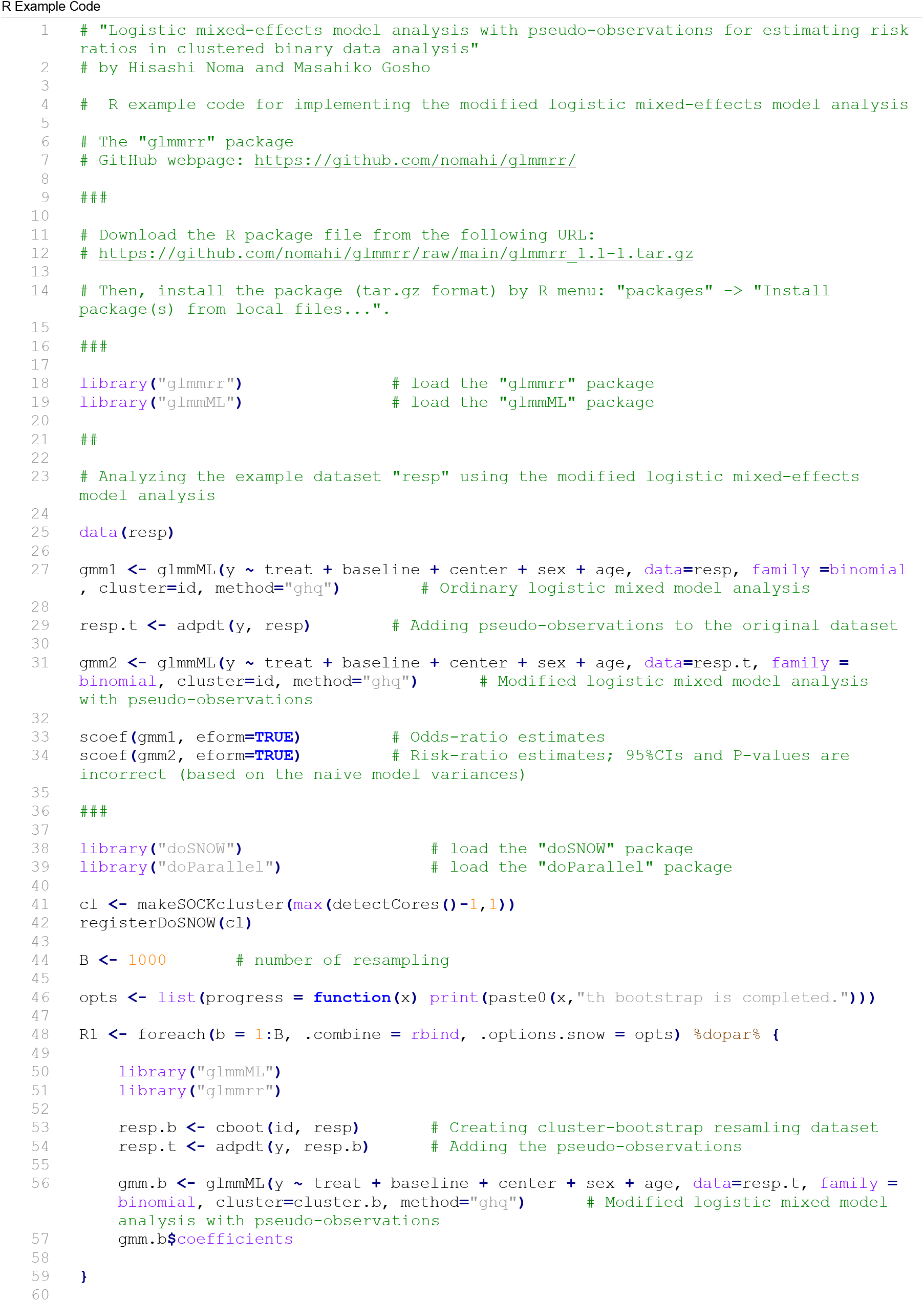
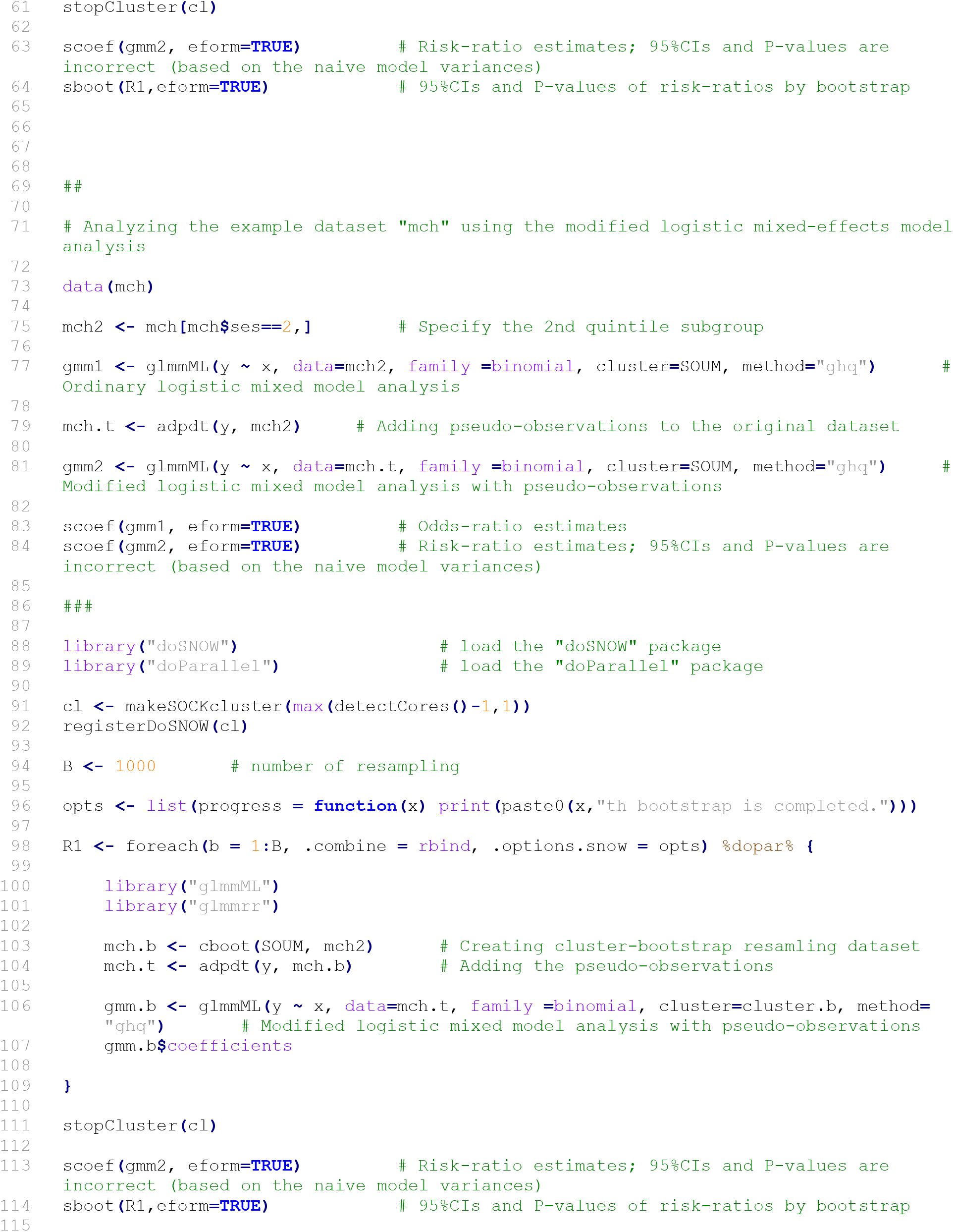

